# Determinants of primaquine and carboxyprimaquine exposures in children and adults with *Plasmodium vivax* malaria

**DOI:** 10.1101/2021.07.05.21259877

**Authors:** Cindy S Chu, James A Watson, Aung Pyae Phyo, Htun Htun Win, Widi Yotyingaphiram, Suradet Thinraow, Nay Lin Soe, Aye Aye Aung, Pornpimon Wilaisrisak, Kanokpich Puaprasert, Mallika Imwong, Warunee Hanpithakpong, Daniel Blessborn, Joel Tarning, Stéphane Proux, Clare Ling, François H Nosten, Nicholas J White

## Abstract

**Background:** Primaquine is the only widely available drug for radical cure of *Plasmodium vivax* malaria. There is uncertainty whether the pharmacokinetic properties of primaquine are altered significantly in childhood or not.

**Methods:** Glucose-6-phosphate dehydrogenase normal patients with uncomplicated *P. vivax* malaria were randomized to receive either chloroquine (25mg base/kg) or dihydroartemisinin-piperaquine (dihydroartemisinin 7mg/kg and piperaquine 55mg/kg) plus primaquine; given either as 0.5 mg base/kg/day for 14 days or 1 mg/kg/day for 7 days. Pre-dose day 7 venous plasma concentrations of chloroquine, desethylchloroquine, piperaquine, primaquine and carboxyprimaquine were measured. Methemoglobin levels were measured either daily or on days 1, 3, 6 and 13, and additionally on day 10 in the primaquine 14-day groups.

**Results:** Day 7 primaquine and carboxyprimaquine concentrations were available for 641 patients. After adjustment for the primaquine mg/kg daily dose, day of sampling, partner drug, and fever clearance, there was a significant non-linear relationship between age and trough primaquine and carboxyprimaquine concentrations, and day methemoglobin levels. Compared to adults 30 years of age, children 5 years of age had trough primaquine concentrations 0.53 (95% CI: 0.39-0.73) fold lower, trough carboxyprimaquine concentrations 0.45 (95% CI: 0.35-0.55) fold lower, and day 7 methemoglobin levels 0.87 (95% CI: 0.58-1.27) fold lower. Increasing concentrations of piperaquine and chloroquine and poor metabolizer *CYP 2D6* alleles were associated with higher day 7 primaquine and carboxyprimaquine concentrations. Higher blood methemoglobin concentrations were associated with a lower risk of recurrence.

**Conclusion:** Young children have lower primaquine and carboxyprimaquine exposures, and lower levels of methemoglobinemia, than adults. Young children may need higher weight adjusted primaquine doses than adults.

## Introduction

Primaquine is used for radical cure of *Plasmodium vivax* and *Plasmodium ovale* malaria, for antimalarial chemoprophylaxis and as a single dose gametocytocide in *Plasmodium falciparum* malaria (1). Although the slowly eliminated 8-aminoquinoline tafenoquine has been registered in a few countries for radical cure or chemoprophylaxis in adults, and pamaquine and quinocide were once used, primaquine is by far the most widely deployed drug in this class. It is generally agreed that the dosing of antimalarial chemotherapy should be designed to provide equivalent exposures to the active antimalarial across the age range (1). There is uncertainty whether or not children have lower exposures to primaquine and its bioactive metabolites than adults for the same weight adjusted doses, and thus whether they should receive higher doses than currently recommended (2–5). Primaquine is combined with either an artemisinin combination treatment (ACT) or chloroquine to prevent relapse of vivax or ovale malaria (radical cure) (1). Primaquine is a pro-drug requiring metabolism to bioactive metabolites mainly via the liver cytochrome P450 isoenzyme *CYP 2D6* (6). The prevalence of intermediate and non-functioning *CYP 2D6* alleles is over 50% in some populations, particularly in Asians (7–9). Decreased *CYP 2D6* function, resulting from intermediate or poor metabolizer genotypes, potentially lowers radical cure efficacy (10, 11).

Chloroquine has been the first-line treatment for the blood stages of *P. vivax* malaria for over 70 years (1). High grade resistance in *P. vivax* remains confined to Oceania and Indonesia (12) but, in the last decade, low grade chloroquine resistance, manifest by earlier appearance of relapses, has been reported increasingly in other locations. Piperaquine (a slowly eliminated bisquinoline), combined with dihydroartemisinin, is a well-tolerated and highly efficacious ACT which provides an excellent alternative blood stage treatment for *P. vivax* malaria (13). An open label two-way randomized controlled trial, conducted on the Thailand-Myanmar border, showed previously that a 7-day high dose primaquine regimen (1 mg/kg/day), combined either with chloroquine or dihydroartemisinin-piperaquine, was non-inferior to the standard 14-day (0.5 mg/kg/day) regimen (14). This large study allowed investigation of the pharmacokinetic properties of primaquine in adults and children, characterization of the determinants of drug exposures, and correlation with therapeutic outcomes.

## Methods

### Clinical trial

This study was conducted by the Shoklo Malaria Research Unit (SMRU), which is located on the northwest Thailand-Myanmar border. Full details of the study have been reported previously (14). In brief, patients who presented to an outpatient clinic with microscopy confirmed uncomplicated *P. vivax* mono-infections were enrolled if they were ≥ 6 months old, and if they were glucose-6-phosphate dehydrogenase (G6PD) normal by the fluorescent spot test. Patients were excluded if they were pregnant or breastfeeding an infant ≤ 6 months, had a hematocrit ≤25%, or had received a blood transfusion within 3 months. Written informed consent was obtained from patients or from parents or carers of children below 18 years old.

Patients were randomized 1:1:1:1 to each of the following groups:

1. Chloroquine (10 mg base/kg on the first two days of treatment, then 5 mg base/kg on day 3; Remedica, Ltd., Cyprus) and primaquine (1 mg base/kg/day for 7 days; Government Pharmaceutical Organization, Thailand)
2. Chloroquine as above and primaquine (0.5 mg base/kg/day for 14 days)
3. Dihydroartemisinin-piperaquine (dihydroartemisinin 7 mg/kg and piperaquine 55 mg/kg for 3 days; Guilin Pharmaceutical company, China) and primaquine (1 mg base/kg/day for 7 days)
4. Dihydroartemisinin-piperaquine as above plus primaquine (0.5 mg base/kg/day for 14 days)

Food and drink were given before drug administration. Primaquine was usually started on the same day as the schizonticide (i.e. the first day of treatment) but in a subset of patients it was started on the third day of treatment. Patients were assessed daily while on treatment. Methemoglobin was measured either daily or on days 1, 3, 6 and 13, and additionally on day 10 in the primaquine 14-day groups, using a transcutaneous pulse oximeter (Masimo Radical-7®). One recruitment clinic did not have access to a transcutaneous oximeter, so 80 study patients did not have methemoglobin measurements. All drug doses were supervised.

A venous blood sample for antimalarial drug levels was taken before drug administration on day 6 (i.e. the 7^th^ day of treatment) +/- 2 days, and plasma was separated and stored at -70°C. Patients were followed at weeks 2 and 4, and then every 4 weeks until 52 weeks. If there was a *P. vivax* recurrence then a venous plasma blood sample for drug levels was taken again. Standard high dose primaquine (0.5 mg base/kg/day for 14 days) and chloroquine were given for the treatment of recurrences. Follow-up was re-started as if newly recruited, and day 6 sampling for drug levels was collected again. The total study duration was 52 weeks from enrolment.

### Drug measurements

Chloroquine/desethylchloroquine, piperaquine, and primaquine/carboxyprimaquine plasma concentrations were measured using three validated liquid chromatography (LC) – tandem mass spectrometry (MS/MS) methods (15–17). The lower limits of quantification (LLOQ) were set to 1.4, 1.5, 1.14 and 4.88 ng/mL for chloroquine/desethylchloroquine, piperaquine, primaquine and carboxyprimaquine, respectively. All three quantification methods were validated according to regulatory standards and three levels of quality control samples were analyzed in triplicate within each batch of clinical samples. Total imprecision (i.e. relative standard deviation) for all quality control samples was below 10% during drug quantification.

### *CYP 2D6* genotyping

We carried out a nested case-control genotyping study to assess the effect of *CYP 2D6* polymorphisms on recurrent vivax malaria. Cases were all patients with a recorded recurrent episode of vivax malaria during follow-up and controls were patients without observed recurrences matched by age and sex (in total n=158 patients). The *CYP 2D6* gene duplications/multiplications and gene deletions were determined by extra-long range polymerase chain reaction (XL-PCR) using a previously published protocol (18). The functional *CYP 2D6* and nonfunctional *CYP2D8* and *CYP2D7* genes were then differentiated by intron 2 sequencing (INT2). The variants *1, *2, *4, *5, *10, *39, and *36 were identified with rapid identification of four polymorphic loci by allele-specific oligonucleotide probes using real-time SNPs genotyping. From the *CYP 2D6* genotyping result the *CYP 2D6* “activity score” was obtained as described previously (15, 19–21).

### Ethics approval

This study was approved by both the Mahidol University Faculty of Tropical Medicine Ethics Committee (MUTM 2011-043, TMEC 11-008) and the Oxford Tropical Research Ethics Committee (OXTREC 17-11) and was registered at ClinicalTrials.gov (NCT01640574).

### Statistical analysis

All drug concentration data were analyzed on the log base 10 scale. Three main generalized additive linear regression models were fitted to the log_10_ primaquine day 7 concentrations; the log_10_ carboxyprimaquine concentrations; and the log_10_ carboxyprimaquine to primaquine ratios. We used the following predictors age (years), mg/kg daily dose of primaquine (the administered mg/kg dose not the target dose), study arm (DHA-piperaquine versus chloroquine), fever clearance time (days), the number of days since start of primaquine dosing (6 versus 4), and the time since the previous primaquine dose (in hours). We excluded drug measurements taken outside of a 18-30 hour window after the previous dose (only trough levels approximately one day after dosing), except for those for which the previous dose was given less than 30 minutes before the drug measurement. More than 90% of samples were taken in this window (Supplementary Figure 1). A smooth spline term with 3 degrees of freedom was used to estimate non-linear effects of age. Individual random intercept terms were specified to account for multiple day 7 measurements in the same patient. To assess any residual effect of *CYP 2D6* diplotypes, we fitted linear regression models to the model residual concentrations with the *CYP 2D6* activity score as a linear predictor. To estimate the effect of the partner drug concentrations (not available for every day 7 measurement), we fitted separate generalized additive regression models to the primaquine and carboxyprimaquine log_10_ concentrations with either the log_10_ chloroquine+desethychloroquine (assuming equal antimalarial activities of parent drug and metabolite and designated as ‘chloroquine’ in the remainder of the paper) concentration or the log_10_ piperaquine concentration as additional predictors.

We fitted a generalized additive regression model to the day 7 blood methemoglobin concentrations (expressed as a percentage of the hemoglobin concentration) with age, mg/kg daily dose of primaquine, study arm (DHA-piperaquine versus chloroquine), fever clearance time (days), and days since start of primaquine dosing as predictors. A spline term with 3 degrees of freedom was fit to age. Secondary models included the *CYP 2D6* activity score (available only for the subset of patients in the nested case control investigation) as an additional covariate, both on the linear scale and as a binary covariate (≤0.5 versus >0.5). The activity score has an arbitrary scale so it is unclear how best to encode it in regression models (i.e. an activity score of 2 versus 1 is not the same as 1 versus 0).

We fitted Cox proportional hazard models to the time to first recurrence (right censored event for patients who did not have a recurrence during follow-up), with continuous predictors: age, partner schizonticidal drug (chloroquine versus piperaquine), and either the log_10_ primaquine or the log_10_ carboxyprimaquine day 7 concentration (they are highly co-linear so we fitted two separate models). Additional models included the day 7 methemoglobin levels as an additional predictor (measured in all sites except the one where a transcutaneous pulse oximeter was not available).

The generalized additive models were fitted in R version 4.0.2 using the package *mgcv* version 1.8. The Cox proportional hazard model used the *survival* package version 3.2. Code in the form of an RMarkdown script along with all analyzed data are available on an accompanying github repository for full reproducibility (https://github.com/jwatowatson/Primaquine_day7).

## Results

Between February 2012 and July 2014, 680 patients with uncomplicated acute *P. vivax* malaria were enrolled. In total 641 patients had at least one day 7 plasma primaquine and carboxyprimaquine concentration measured, either during the enrollment episode or during a recurrent *P. vivax* episode (there were a total of 720 episodes with day 7 concentrations including patients with multiple episodes). For 692 of these day 7 concentrations, the corresponding plasma chloroquine or piperaquine concentration was also available (measured from the same blood sample). We excluded 41 drug measurements considered not to be trough levels (see Methods) resulting in a total of 679 day 7 measurements included in the analysis.

In total, 71 of the patients with measured primaquine and carboxyprimaquine concentrations were 6 to ≤10 years of age (median weight 20kg, range 13-30kg), and 34 were ≤5 years of age (median weight 14kg, range 8-20kg) (Table 1). Three patients were less than 3 years old, and the youngest was 1 year 6 months old. Fever and parasite clearance times were similar across all age groups. The proportion of patients with microscopy detected gametocytemia was higher in the older age groups >10 years.

**Table 1.**
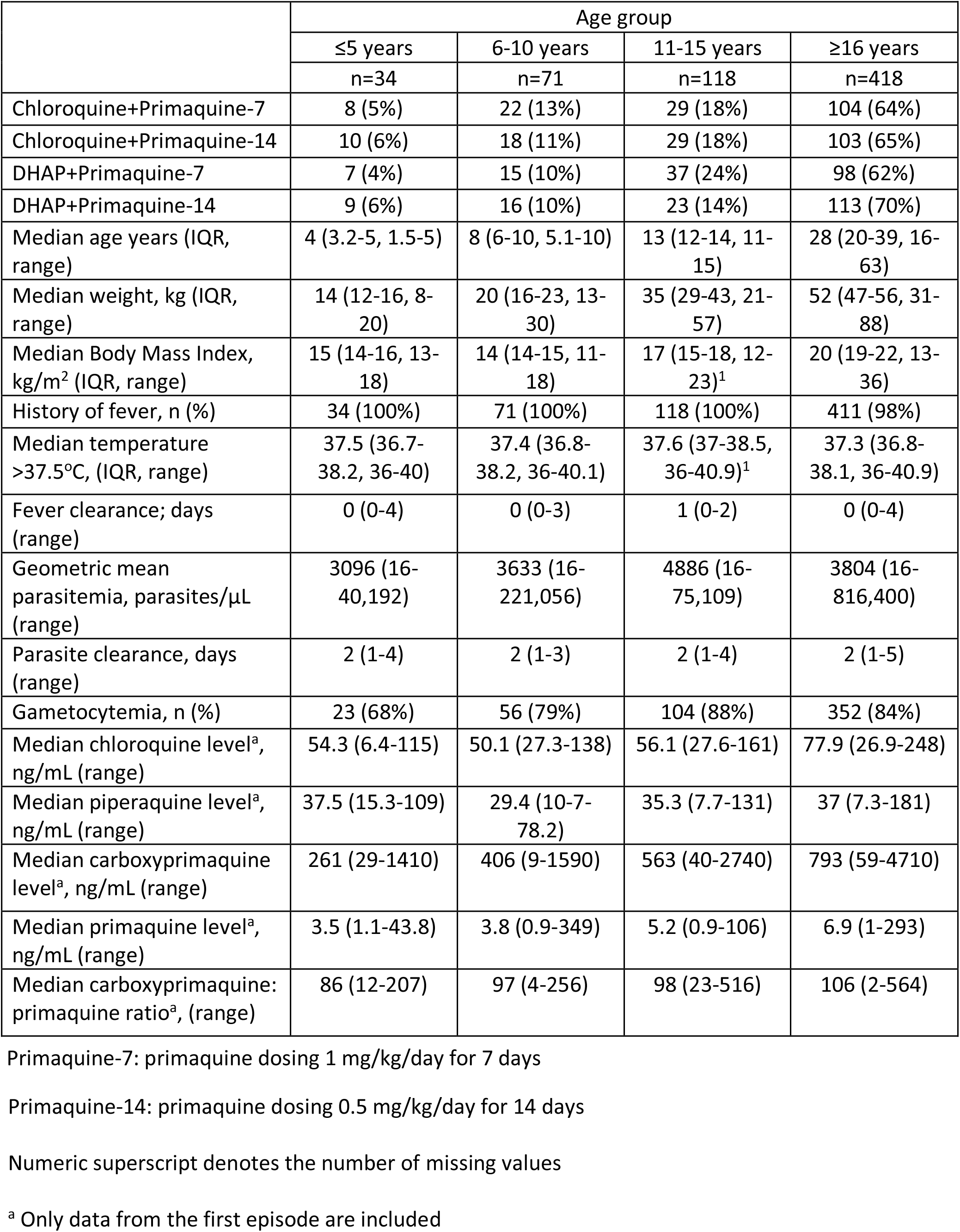
Patient characteristics at enrolment by age group

### Effect of age

After adjustment for the daily mg/kg primaquine dose, the number of days of dosing, the fever clearance times, and the partner drug (piperaquine versus chloroquine), there was a significant non-linear association between age and day 7 primaquine trough concentration (*p*<10^−16^ for the smooth spline term fit to age) and day 7 carboxyprimaquine trough concentration (*p*<10^−16^ for the smooth spline term fit to age) (Figure 1). For both primaquine and carboxyprimaquine, increasing age correlated with higher trough levels. The strongest effect was seen for the carboxyprimaquine trough levels. Thus the ratio of carboxyprimaquine to primaquine was also associated with age, with an approximate doubling when comparing patients aged ≤5 years versus patients 30 years or older (Figure 1).

**Figure 1:**
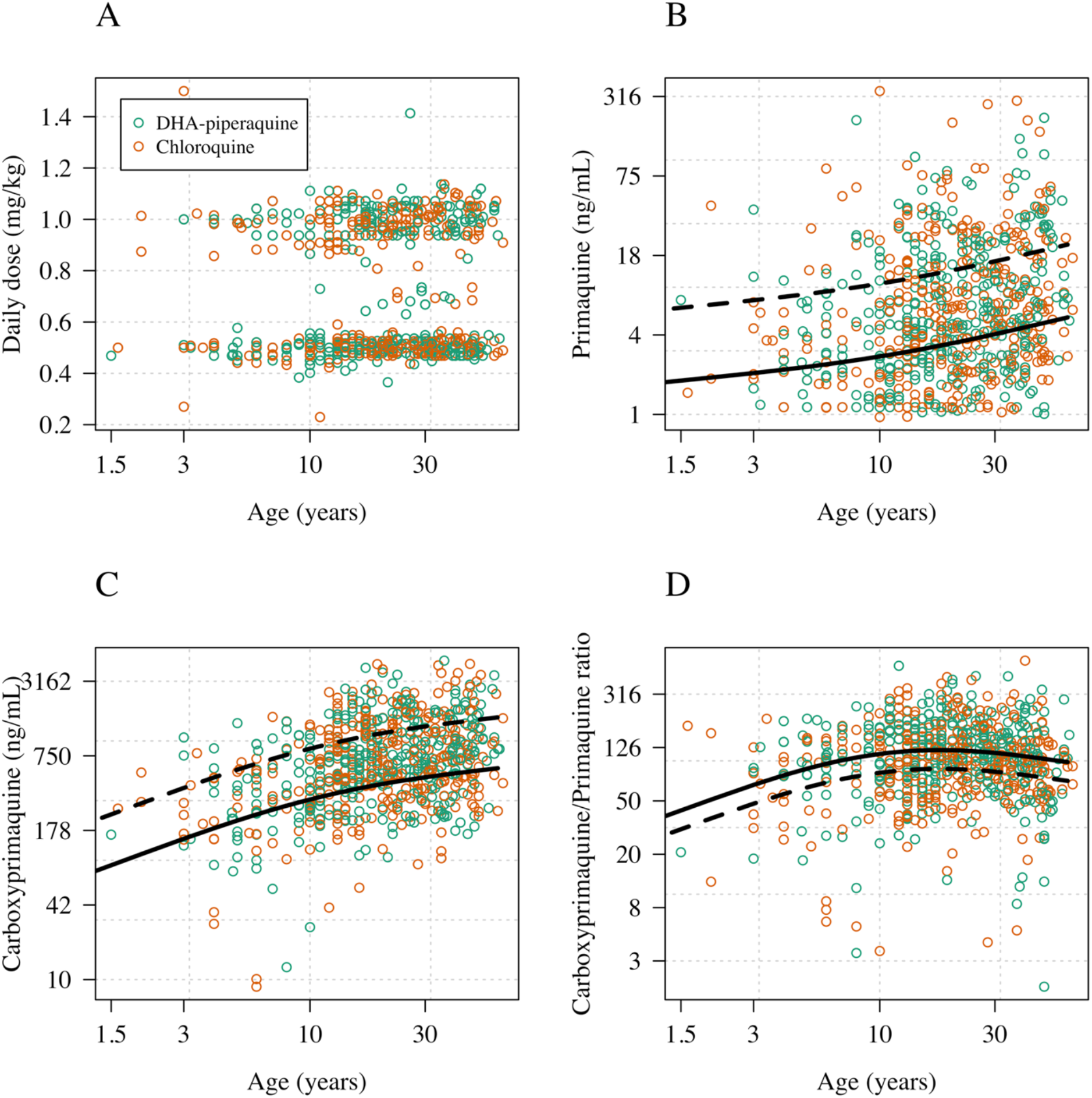
The effect of age on primaquine and carboxyprimaquine exposures in patients with acute vivax malaria. Panel A: age versus mg/kg primaquine daily dose; Panel B: age versus the primaquine concentration on day 7; Panel C: age versus the carboxyprimaquine concentration on day 7; Panel D: age versus the ratio of the day 7 carboxyprimaquine to primaquine concentration. The black lines show model fits using a generalized additive model with a smooth spline term for age and adjusting for daily mg/kg dose; number of days of primaquine received, time since last dose, partner drug and fever clearance time (dashed: mean predicted concentration for 1mg/kg daily; solid: mean predicted concentration for 0.5 mg/kg daily).

### Effect of partner drug

Our previously reported healthy volunteer studies showed that co-administration of piperaquine and chloroquine both increased exposures to primaquine and carboxyprimaquine (15, 16). In this study of patients with vivax malaria, a ten-fold increase in the chloroquine day 7 concentration was associated with a 2.5 fold increase in day 7 primaquine concentrations (95% CI: 1.9 to 3.1; *p*=0.0003), and a 2.0 fold increase in day 7 carboxyprimaquine concentrations (95% CI: 1.7 to 2.4; *p*=0.0002). A ten-fold increase in the piperaquine day 7 concentrations was associated with a 2.5 fold increase in primaquine concentrations (95% CI: 1.9 to 3.2; p=0.0005), and a 2.5 fold increase in carboxyprimaquine concentrations on day 7 (95% CI: 2.1 to 3.0; p=10^−7^). Scatterplots showing the univariate relationships between the partner drug concentrations and primaquine/carboxyprimaquine concentrations are shown in Supplementary Figure 2. Treatment with chloroquine or piperaquine had no statistically significant effect on the carboxyprimaquine/primaquine ratios.

### Effect of *CYP 2D6* genotypes

*CYP 2D6* genotypes were determined in 154 patients. The estimated *CYP 2D6* allele frequencies in this population were as follows: 25% for *1; 23% for *2; 2% for *4; 10% for *5; 38% for *10; 0.5% for *39; and 1% for *36. Alleles *4, *5, and *36 are considered null-metabolizer alleles (enzyme activity score of 0); *10 is a poor metabolizer allele (activity score of 0.25); and *1 and *39 are normal metabolizer alleles (activity score of 1). In total, there were 3 patients with an activity score of 0 (non-metabolizers) and 36 patients with activity scores of either 0.25 or 0.5 (poor metabolizers). For primaquine day 7 plasma concentrations, model residuals (defined as the observed concentration minus the predicted concentration, both on the log scale, where the model is based on age, mg/kg dose, fever clearance, time since last dose, number of days of dosing, and partner drug), were correlated significantly with the genotype-defined *CYP 2D6* enzyme activity scores (Figure 2; ρ;= -0.2, p=0.004). Patients with low *CYP 2D6* activity scores had, on average, higher observed concentrations of primaquine than predicted (i.e. positive residuals), whereas patients with normal activity scores had lower concentrations than expected (i.e. negative residuals). For plasma carboxyprimaquine concentrations, a trend in the same direction was observed, but the correlation was substantially smaller, and it was not significant (p=0.1).

**Figure 2:**
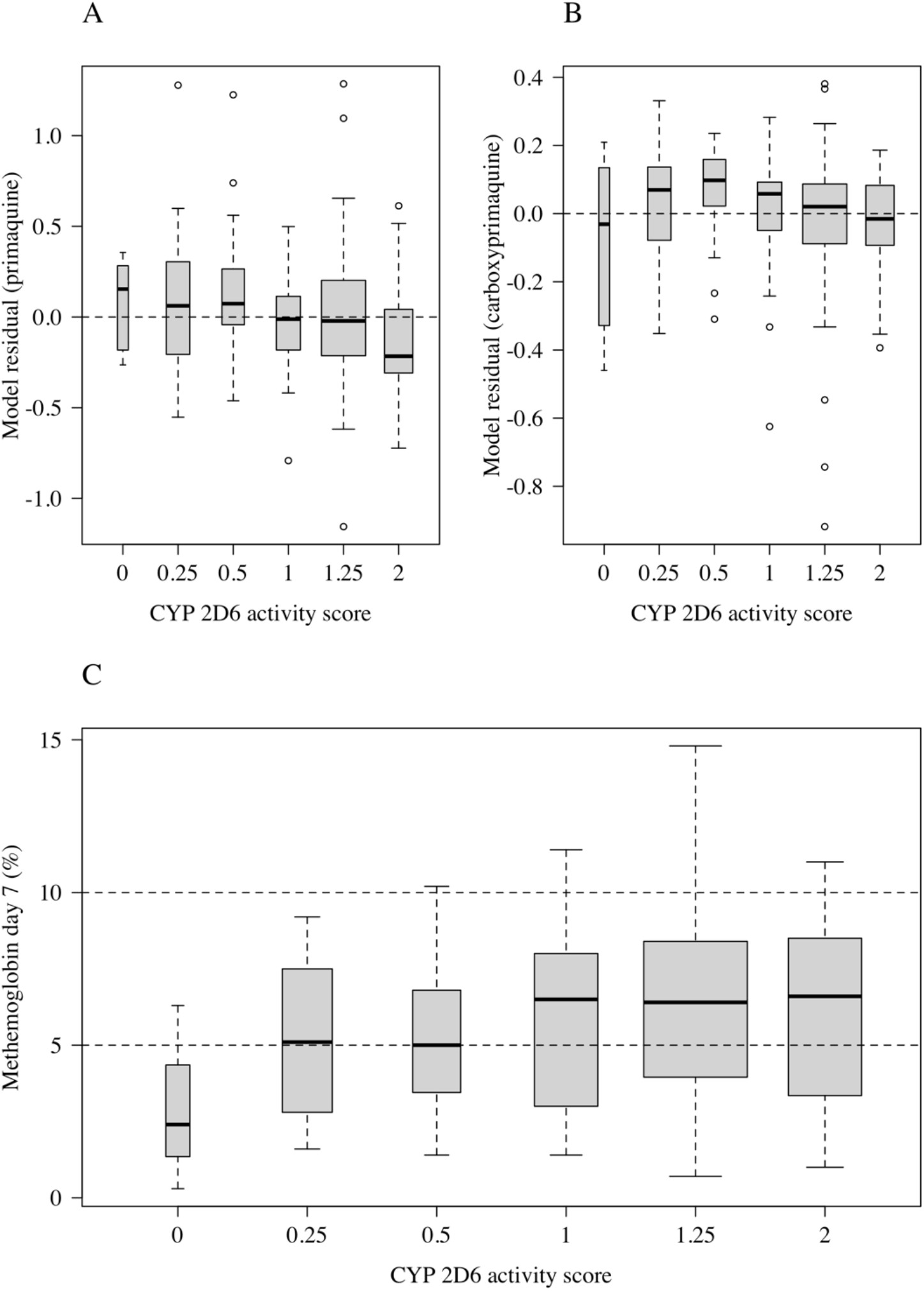
The relationship between *CYP 2D6* genotypes and primaquine and carboxyprimaquine exposure, and day 7 methemoglobin levels. In panels A and B, the y-axis shows the model residuals (observed minus predicted) for the main model of exposure (adjusting for age, daily mg/kg primaquine dose, partner drug, and fever clearance time). In panel C, the y-axis shows the observed day 7 methemoglobin %. In all three panels the x-axis is the activity score predicted from the *CYP 2D6* diplotype (20). Each box-and-whisker plot shows the median value, the interquartile range and the range. The box widths are proportional to the square-roots of the number of observations per activity score.

### Methemoglobinemia and hemolysis

Primaquine causes predictable methemoglobinemia. As reported previously methemoglobin concentrations rose over the first week of primaquine treatment, and then plateaued in the 14-day primaquine groups. The day 7 values were usually the peak recorded values (Supplementary Figure 3). Methemoglobin (expressed as a percentage of the hemoglobin concentration) was lower in children than in young adults (*p*<10^−16^ for a spline term fit to age in a generalized additive regression model, Figure 3A), and was lower in patients with loss of function *CYP 2D6* polymorphisms (Figure 2C; absolute decrease of 1.2 percentage points in the mean day 7 methemoglobin level in patients with an activity score ≤ 0.5 compared to patients with an activity score > 0.5; 95% CI: 0.1-2.3%, *p*=0.04). Day 7 methemoglobin levels were associated with both primaquine and carboxyprimaquine day 7 plasma concentrations. A ten-fold increase in primaquine day 7 concentrations was associated with an absolute increase of 0.7 percentage points in day 7 methemoglobin levels (95% CI: 0.14-1.23), and a ten-fold increase in carboxyprimaquine concentrations was associated with an absolute increase of 1.1 percentage points in day 7 methemoglobin levels (95% CI: 0.5 – 2.0). Four patients stopped taking primaquine because of elevated methemoglobin levels and associated symptoms. Two of these patients stopped on day 5 and, as the day 6 primaquine dose was not administered, they had low day 7 primaquine and carboxyprimaquine levels. One of these patients had a recorded methemoglobin level of 20.9% on day 6 followed by 18.1% on day 7. The other three patients had peri-oral cyanosis (blue lips). Their peak recorded methemoglobin levels were between 10 and 12%.

**Figure 3:**
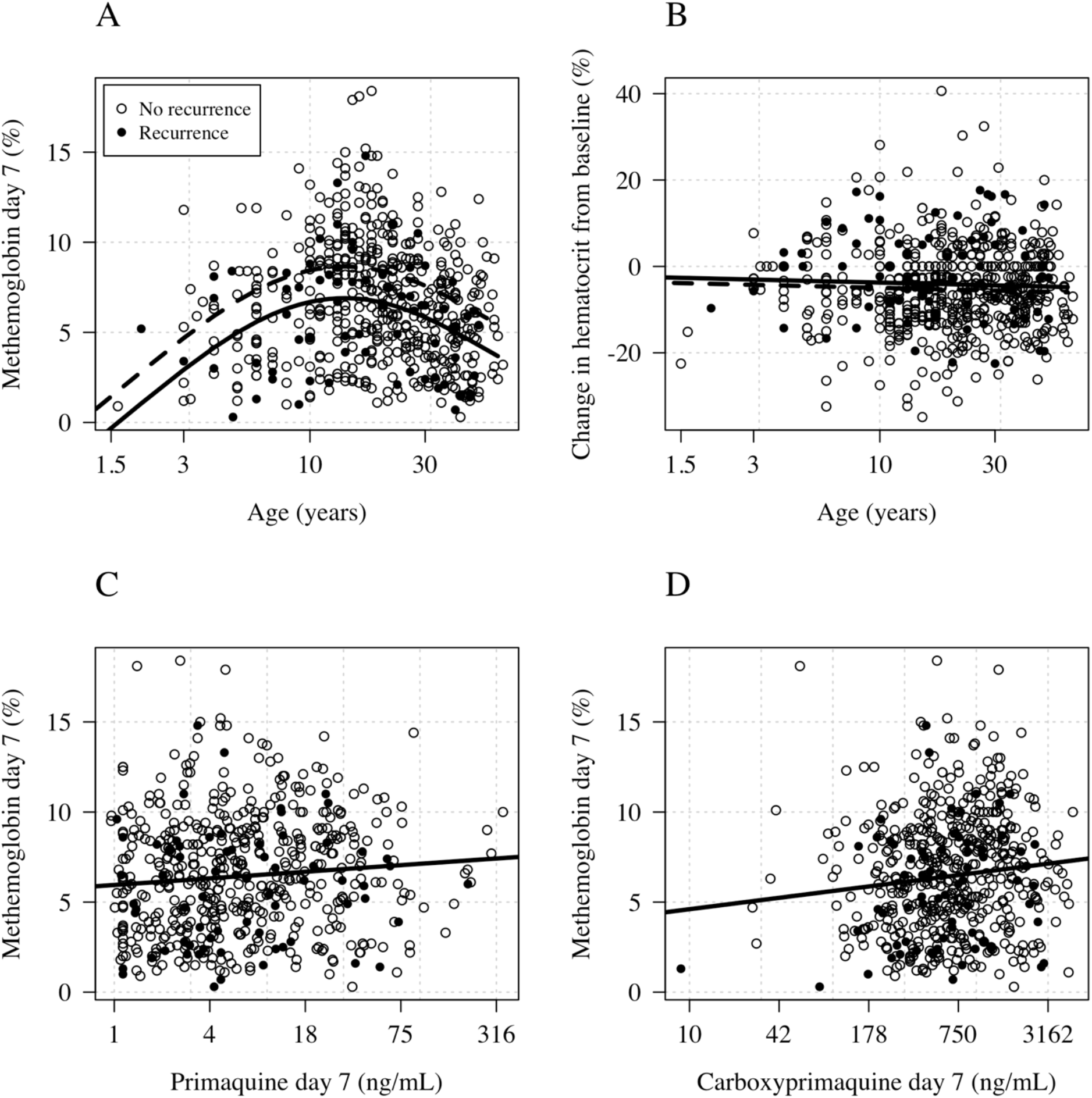
Age and drug exposure relationships with hemolysis and methemoglobinemia. Panel A: Age and day 7 methemoglobin Panel B: Age and the day 7 change in hematocrit from baseline. Panel C: Day 7 methemoglobin and pre-dose primaquine concentrations. Panel D: Day 7 methemoglobin and pre-dose carboxyprimaquine concentrations. In panels A&B the black lines show model fits under a generalized additive model with a smooth spline term for age and adjusting for daily mg/kg dose; number of days of primaquine received, the time since last dose, and G6PD deficiency (dashed: mean predicted concentration for 1mg/kg daily; solid: mean predicted concentration for 0.5 mg/kg daily).

There was no relationship between age and hemolysis defined as the percentage drop in hematocrit between baseline and day 7 (Figure 3B).

### Primaquine exposure and *P. vivax* recurrence

We did an exploratory analysis of the relationship between primaquine exposures and times to the first *P. vivax* recurrence. A total of 564 patients had recorded day 7 drug levels and methemoglobin levels, and 70 had at least one recurrence during follow-up. After adjusting for age and partner drug, day 7 concentrations of primaquine and carboxyprimaquine were not associated with the risk of recurrence, but a 1% absolute increase in day 7 methemoglobin was associated with a hazard ratio for recurrence of 0.9 (95% CI: 0.85-0.99, p=0.02). Two of the three patients with a *CYP 2D6* activity score of 0 had a *P. vivax* recurrence, and one of them had 2 recurrences (the interval between recurrences was >65 days). Studies are ongoing to characterize the relationship between *CYP 2D6* genotypes and risk of *P. vivax* relapse in this population.

## Discussion

Radical cure of *P. vivax* malaria is necessary to prevent relapses. Relapses of vivax malaria are a major cause of morbidity and developmental delay in children living in endemic areas, so optimization of primaquine dosing is critical in this age group. The higher *P. vivax* relapse rates in children compared with adults living in endemic areas observed in some studies (22) are attributed usually to lower levels of immunity, but lower drug exposures could also be a contributory factor. For many antimalarial drugs, exposures in children are lower than in adults and an increase in the weight adjusted dosing is needed (1). The limited pharmacokinetic evidence to date for primaquine does not provide a clear picture, with a study from Papua New Guinea suggesting little difference between adults and older children (2), whereas a study of single dose primaquine (used as a *P. falciparum* gametocytocide) in Tanzania (3) and a study of primaquine for *P. vivax* radical cure in Brazil (4) both suggesting that younger children had lower exposures. The relationship between plasma primaquine concentrations and antimalarial effect is complex. Primaquine is metabolized rapidly (elimination half-life ∼5 hours) via monoamine oxidase to a biologically inert metabolite carboxyprimaquine and, through a separate pathway, mainly via *CYP 2D6*, to several active hydroxylated metabolites (6, 23). The exact mechanism of antimalarial action remains uncertain. Recent laboratory studies suggest that these unstable hydroxylated metabolites are oxidized to quinoneimines generating local hydrogen peroxide - which is parasiticidal. The quinoneimines in turn are substrates for cytochrome P450 NADPH:oxidoreductase (POR or CPR) resulting in hydrogen peroxide accumulation and augmenting the parasiticidal effect (24). These reactions may mediate both the antimalarial therapeutic effects, and also the oxidant hemolytic adverse effects. Alternatively the quinoneimine itself may cause the hemolytic and possibly parasiticidal effects by forming irreversible ligands with heme. There is also evidence for synergy between the 8-aminoquinolines and other quinoline antimalarials in liver stage and blood stage activities (25, 26). The 8-aminoquinolines have innate schizonticidal activity as well as radical curative activity and 4-aminoquinoline antimalarials increase primaquine concentrations (15, 16, 26). Unfortunately, although the metabolic pathway to the production of the active primaquine metabolites has been characterized ex-vivo, no validated measure of their concentrations, or of their more stable metabolites, in whole blood, plasma, or urine has yet been correlated with antimalarial therapeutic responses in-vivo. Thus, measurement of primaquine and carboxyprimaquine concentrations does not provide a direct correlate of antimalarial activity. Furthermore, because the apparent volume of distribution of carboxyprimaquine is not known, the proportion of parent drug that passes through this inactive metabolic pathway cannot be estimated.

This large study of day 7 “trough” primaquine and carboxyprimaquine plasma concentrations shows clearly that exposures to both parent compound and inactive metabolite are lower in children than in adults. This is unlikely to be explained by lower oral bioavailability as primaquine is generally well absorbed, the children in the study were clinically well by seven days after starting treatment (14), all doses were observed, and only 7 children ≤10 years old (≤27kg) vomited. It presumably reflects either larger primaquine (and carboxyprimaquine) distribution volumes, or increased clearance, or both. The lower ratio of carboxyprimaquine to primaquine in children may also be explained by these age-related pharmacokinetic changes. Although greater diversion down the bioactivating CYP pathway cannot be excluded as an explanation for these findings-there is no convincing evidence for augmented drug efficacy (e.g. no relationship with fever or parasite clearance times) or increased hemolytic toxicity in children to support this. Furthermore, in this study, methemoglobinemia was lower in children than in young adults, indicating that formation of oxidant metabolites was reduced (27). Lower methemoglobin concentrations were associated with an increased risk of *P. vivax* recurrence. The importance of *CYP 2D6* primaquine bioactivation in the generation of oxidant metabolites was evidenced by the lower levels of methemoglobinemia in patients with loss of function *CYP 2D6* polymorphisms (Figure 2C). Taken together this evidence strongly suggests that children have lower exposures to the biologically active metabolites of primaquine than do adults receiving the same weight adjusted doses.

This study was sufficiently large that the influence of other covariates affecting primaquine exposures could be examined. We have reported previously that co-administration of quinoline or structurally related antimalarials increases primaquine and carboxyprimaquine concentrations (15, 16). This was confirmed in this study. The effects on the parent compound of chloroquine and piperaquine were similar whereas piperaquine had a larger effect on carboxyprimaquine. In Asia loss of function *CYP 2D6* genetic polymorphisms (mainly the *10 allele) are common. In this study low *CYP 2D6* activity scores (derived from the genotype) were associated with higher primaquine levels and lower levels of methemoglobinemia as would be expected from reduced metabolic conversion through this bioactivation pathway.

There are several limitations to this study. This analysis is limited to drug concentrations at one time point during radical cure treatment providing a limited description of drug exposures. Very young children are under-represented, and the analysis does not include any infants < 1 year old. In addition, *CYP 2D6* genotyping was not performed in all patients.

There is no evidence that the adverse effects of primaquine are significantly worse in children than adults. The main therapeutic implication of this large pharmacometric evaluation is that children may require larger weight adjusted doses of primaquine than adults.

## Data Availability

All data are available on the linked github repo
https://github.com/jwatowatson/Primaquine_day7

## Acknowledgements

This research was funded by The Wellcome Trust. A CC BY or equivalent license is applied to the author accepted manuscript arising from this submission, in accordance with the grant’s open access conditions. The authors would like to thank the SMRU staff for their hard work in running this study and the MORU Clinical Pharmacology Laboratory in Bangkok for their diligent work in sample processing and drug measurements. Special appreciation goes to the contributions of Htet Htet Chaw, Nwe Wah Lin, Thida Zin, Say Paw, Naw Eh Shee, Thu Lay Paw, Rattanporn Raksapraidee, Moo Koo Paw, the data entry team headed by Jacher Wiladphaingern, the logistics team, and the laboratory teams headed by Kanlaya Sriprawat and Germana Bancone.

We would also like to thank the Data Safety and Monitoring Board: Bob Taylor, Charles Woodrow, and Professor Nicholas Day for their time and commitment.

## Legends to Figures

**Supplementary Figure 1:**
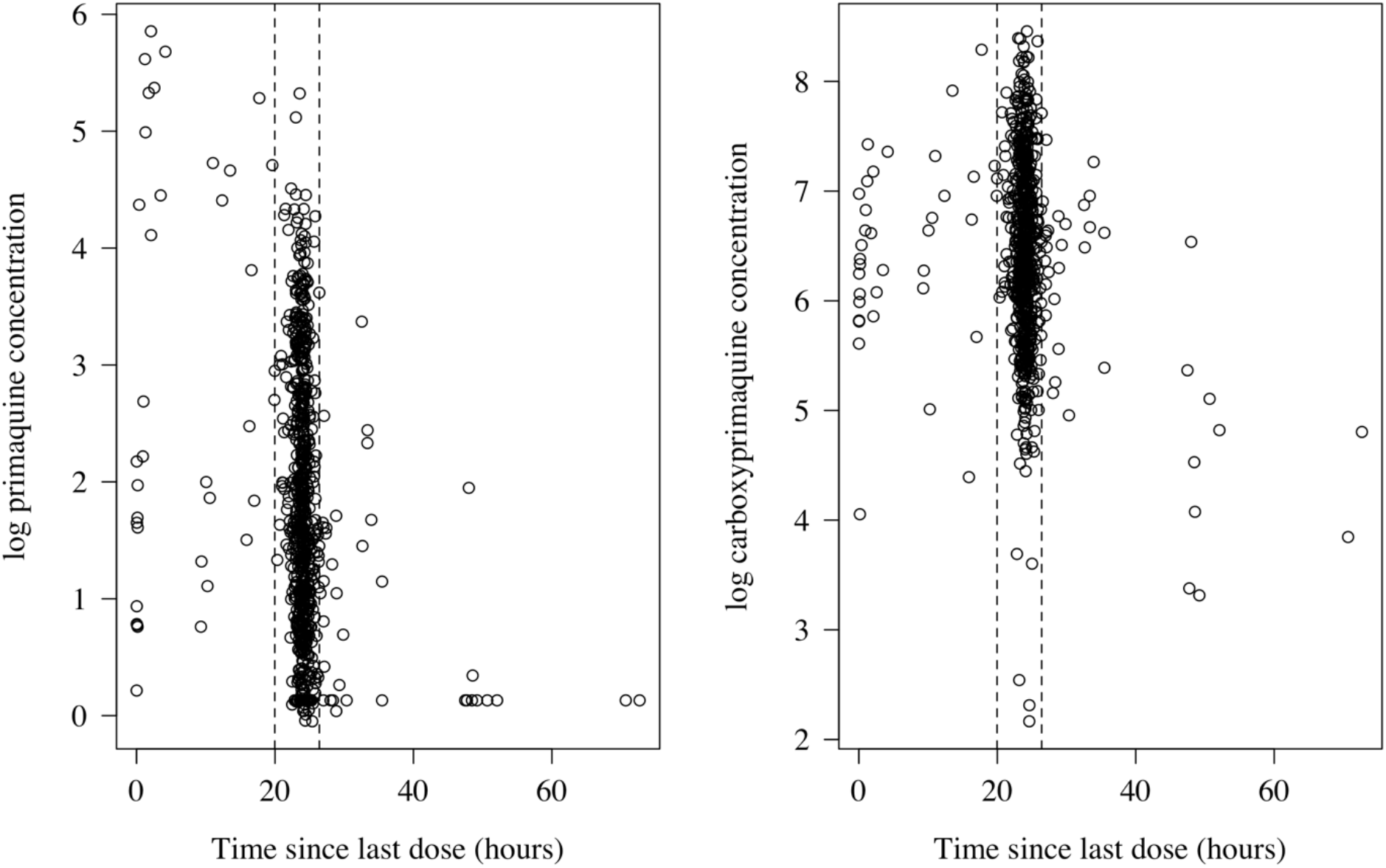
time since previous dose versus the primaquine and carboxyprimaquine blood concentration. The dashed vertical lines show the 5^th^ and 95^th^ quantiles of the time since last dose.

**Supplementary Figure 2:**
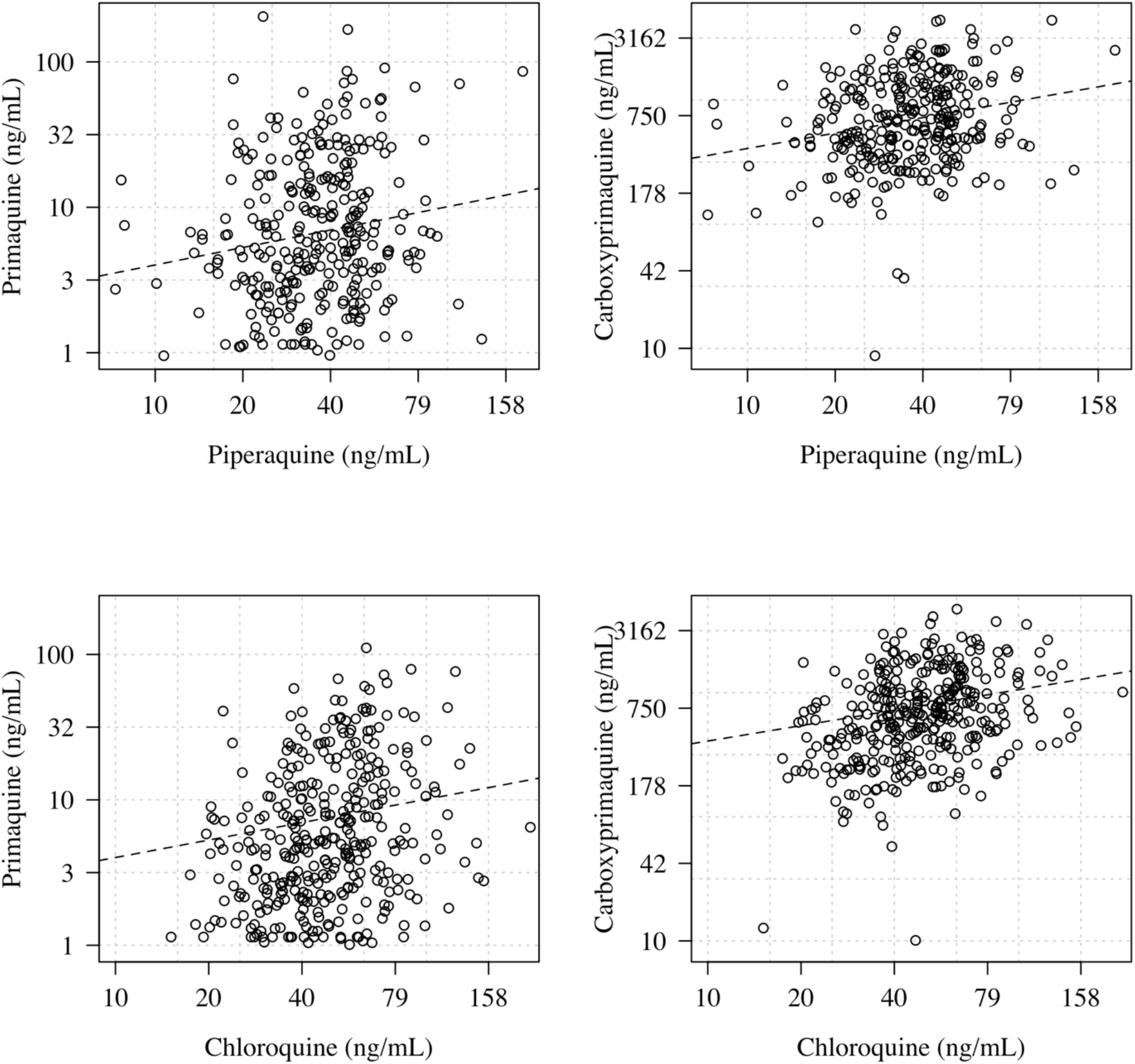
Primaquine and carboxyprimaquine log concentrations versus chloroquine and piperaquine log concentrations measured at the same timepoints. The dashed lines show a univariate linear regression to highlight the trend.

**Supplementary Figure 3:**
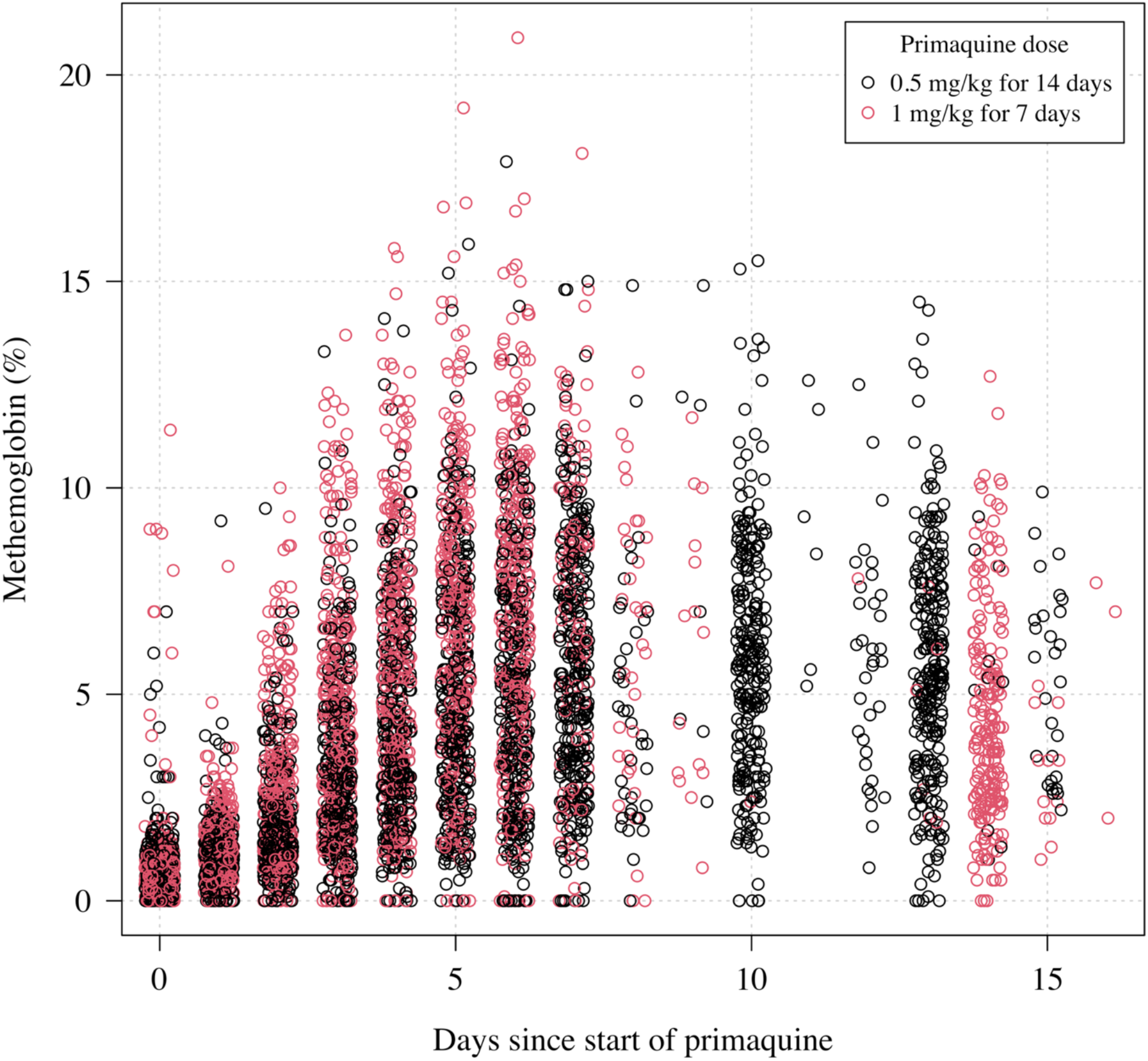
Methemoglobin as function of the time since starting primaquine (black circles: 0.5 mg/kg target dose for 14 days; red circles: 1 mg/kg target dose for 7 days). We added random jitter to the time in days for visual clarity.

**Supplementary Table 1:** Estimated regression coefficients (standard errors) for the linear terms in the main models. NI: not included. Age is included in these models as a non-linear term and so it is excluded from this table (effects are shown in Figures 1 & 3). For the time to recurrence model, the coefficients and standard errors are given on the linear scale (hazard ratio is given by the exponential of the coefficient).

| Outcome variable | Regression type | Dose (mg/kg) | Partner drug | Days of primaquine | Fever clearance (days) | CYP 2D6 activity score < 1 | G6PD deficiency | Methb (%) |
| --- | --- | --- | --- | --- | --- | --- | --- | --- |
| PQ (log ng/mL) | linear | 1.14<br>(0.06) | 0.014<br>(0.03) | 0.038 (0.047) | -0.017<br>(0.021) | NI | NI | NI |
| CPQ (log ng/mL) | linear | 0.85<br>(0.040) | -0.020<br>(0.021) | 0.042 (0.032) | -0.027<br>(0.014) | NI | NI | NI |
| log CPQ/PQ | linear | -0.28<br>(0.043) | -0.039<br>(0.022) | -0.0040<br>(0.035) | -0.0086<br>(0.016) | NI | NI | NI |
| Methb (%) | linear | 3.14<br>(0.92) | NI | -1.11 (0.77) | NI | -1.21<br>(0.59) | -0.43 (0.86) | NI |
| Hb reduction (%) | linear | -2.43<br>(1.43) | -2.90<br>(0.73) | 3.07 (1.13) | -2.10<br>(0.50) | NI | -12.0 (1.67) | NI |
| Time to recurrence | Cox proportional hazards | NI | -0.23<br>(0.24) | NI | NI | NI | NI | -0.09<br>(0.039) |

